# Chronic SARS-CoV-2 infection and viral evolution in a hypogammaglobulinaemic individual

**DOI:** 10.1101/2021.05.31.21257591

**Authors:** Maia Kavanagh Williamson, Fergus Hamilton, Stephanie Hutchings, Hannah M. Pymont, Mark Hackett, David Arnold, Nick A Maskell, Alasdair MacGowan, Mahableshwar Albur, Megan Jenkins, Izak Heys, Francesca Knapper, Mustafa Elsayed, Rachel Milligan, The COVID-19 Genomics UK (COG-UK) Consortium, Peter Muir, Barry Vipond, David A Matthews, Ed Moran, Andrew D. Davidson

## Abstract

There is widespread interest in the capacity for SARS-CoV-2 evolution in the face of selective pressures from host immunity, either naturally acquired post-exposure or from vaccine acquired immunity. Allied to this is the potential for long perm persistent infections within immune compromised individuals to allow a broader range of viral evolution in the face of sub-optimal immune driven selective pressure. Here we report on an immunocompromised individual who is hypogammaglobulinaemic and was persistently infected with SARS-CoV-2 for over 290 days, the longest persistent infection recorded in the literature to date. During this time, nine samples of viral nucleic acid were obtained and analysed by next-generation sequencing. Initially only a single mutation (L179I) was detected in the spike protein relative to the prototypic SARS-CoV-2 Wuhan-Hu-1 isolate, with no further changes identified at day 58. However, by day 155 the spike protein had acquired a further four amino acid changes, namely S255F, S477N, H655Y and D1620A and a two amino acid deletion (ΔH69/ΔV70). Infectious virus was cultured from a nasopharyngeal sample taken on day 155 and next-generation sequencing confirmed that the mutations in the virus mirrored those identified by sequencing of the corresponding swab sample. The isolated virus was susceptible to remdesivir *in vitro*, however a 17-day course of remdesivir started on day 213 had no effect on the viral RT-PCR cycle threshold (C_t_) value. On day 265 the patient was treated with the combination of casirivimab and imdevimab. The patient experienced progressive resolution of all symptoms over the next 8 weeks and by day 311 the virus was no longer detectable by RT-PCR. The ΔH69/ΔV70 deletion in the N-terminus of the spike protein which arose in our patient is also present in the B.1.1.7 variant of concern and has been associated with viral escape mutagenesis after treatment of another immunocompromised patient with convalescent plasma. Our data confirms the significance of this deletion in immunocompromised patients but illustrates it can arise independently of passive antibody transfer, suggesting the deletion may be an enabling mutation that compensates for distant changes in the spike protein that arise under selective pressure.

## Introduction

As the COVID-19 pandemic enters its second year and vaccines are deployed around the world there has been considerable attention paid to the emergence of viral variants with the potential for greater transmissibility (*1-3*), disease severity (*4*) and vaccine evasion (*5*). Such variants may arise through an incremental process as they pass through a population, or potentially within an individual who fails to clear infection promptly. Persistent infection, well beyond the 14 days by which most healthy people are believed to clear the virus (*6*), is increasingly recognised among certain immunocompromised patients, commonly those with B-cell deficiencies (*7*). Immunotherapy with convalescent plasma (CP) has been proposed as a potential therapy (*8, 9*). However, a recent report raises concern that such interventions have the potential to drive the emergence of diverse viral variants capable of evading the antibodies present in CP and by extension, the immune responses generated by currently available vaccines (*10*). Following the administration of two rounds of CP, and thought to be driven by it, a new dominant viral strain arose bearing a two amino acid H69/V70 deletion (ΔH69/ΔV70) in the N terminal domain (NTD) of the spike protein together with a D796H mutation. The authors hypothesised that this deletion compensated for a reduction in infectivity caused by the latter mutation.

In this report we describe a patient with persistent SARS-CoV-2 infection associated with hypogammaglobulinemia and persistent lymphopenia as a result of treatment for chronic lymphocytic leukaemia. Consistently positive by RT-PCR testing, viral viability was confirmed by successful culture 6 months after initial diagnosis. Treatment of the patient with remdesivir failed to clear the virus, which was not resistant to the drug *in vitro*. In contrast, after combined treatment with casirivimab and imdevimab, the virus was undetectable in swab samples 45 days post-treatment and the patient experienced a dramatic clinical improvement. Next generation sequencing (NGS) demonstrated the acquisition of the ΔH69/ΔV70 deletion prior to any therapy, potentially supporting the concept that it may be an enabling mutation, restoring infectivity otherwise impaired by point mutations in the spike protein.

## Methods

### Cell culture

African green monkey kidney (VeroE6, ATCC® CRL 1586™) cells constitutively expressing TMPRSS2 (VeroE6/TMPRSS2 (*11*) obtained from NIBSC, UK) and human Caco-2 cells expressing ACE2 (Caco-2-ACE2; a kind gift from Dr Yohei Yamauchi, University of Bristol) were cultured in Dulbecco’s Minimal Essential Media with GlutaMAX (DMEM, Gibco™, ThermoFisher) containing 10% foetal bovine serum (FBS, Gibco) and 0.1mM non-essential amino acids (NEAA, Sigma Aldrich). Cells were grown at 37 °C in 5% CO_2_.

### Viruses used and isolated

The SARS-CoV-2 isolates SARS-CoV-2/human/Liverpool/REMRQ0001/2020 (*12*) and hCoV-19/England/204690005/2020 (lineage B.1.1.7; GISAID ID: EPI_ISL_693401; kindly provided by Professor Wendy Barclay, Imperial College, London and Professor Maria Zambon, Public Health England) were used in this study. Virus isolation in this study was performed from a nasopharyngeal swab sample collected in virus transport media. The sample was filtered through a 0.2 µm filter and added to VeroE6/TMPRSS2 cells cultured in Eagle’s Minimum Essential Media with GlutaMAX (MEM, Gibco™, ThermoFisher) containing 2% FBS, 0.1mM NEAA, penicillin (100 units/ml) and streptomycin (100 μg/ml). By day 13 virological cytopathic effect (CPE) was evident. Supernatant was collected and passed through a 0.2 µm filter and a viral stock (referred to as hCoV-19/England/BRIS-MKW1/2020) prepared using VeroE6/TMPRSS2 cells. All work with infectious SARS-CoV-2 virus was conducted in a Class III microbiological safety cabinet in a containment level 3 facility at the University of Bristol.

### Quantitation of viral RNA

Laboratory diagnostic testing for all samples received was performed using the Aptima SARS-CoV-2 transcription mediated assay (TMA) on the Panther system (Hologic, USA) followed by semi-quantitative RT-PCR using the Closed NeuMoDx SARS-CoV-2 commercial assay (Qiagen) with automated RNA extraction and concentration, PCR reagent preparation, and nucleic acid amplification/detection. 300 µl of primary sample was pretreated with NeuMoDx Viral Lysis Buffer prior to loading onto the NeuMoDx System. Targets for this assay are regions of the SARS-CoV-2 nucleocapsid (N) and nsp2 genes.

To quantitate virus grown in the laboratory, viral RNA was extracted from infected cell culture supernatants using a QIAamp Viral RNA Isolation kit (Qiagen) according to the manufacturer’s instructions with elution of RNA in 60 µl of AVE solution. 5 µl of each sample was used for qRT-PCR analysis as previously described (*13*). SARS-CoV-2 RNA genome equivalence was determined using RNA extracted from titred virus and a AccuPlexTM SARS-CoV-2 Reference Material Kit (SeraCare, Milford, US) as standards.

### Immunofluorescence infectivity assay

VeroE6/TMPRSS2 cells were seeded for 18 hours in DMEM in µClear 96-well Microplates (Greiner Bio-one). The culture supernatant was removed, and cells were infected with hCoV-19/England/BRIS-MKW1/2020 in duplicate, in a 2-fold dilution series. Following 18 hours incubation, cells were fixed in 4% PFA for 60 minutes, permeabilised, blocked and stained with DAPI (Sigma Aldridge) and antibodies against the SARS-CoV-2 N protein (200-401-A50, Rockland) and dsRNA (10010200, Scicons) followed by appropriate secondary antibodies. Images were acquired on the ImageXpress Pico Automated Cell Imaging System (Molecular Devices) using a 10X objective. Stitched images of 9 fields covering the central 50% of the well were analysed for infected cells using Cell Reporter Xpress software (Molecular Devices). Briefly, cell number was determined by automated counting of DAPI stained nuclei, infected cells were determined as those cells in which positive N staining was detected associated with a nuclei. Data were plotted using GraphPad Prism v8.4.3.

### Direct RNA sequencing

VeroE6/TMPRSS2 cells were seeded into a T75 cell culture flask and infected with hCoV-19/England/BRIS-MKW1/2020 at an MOI of 0.01 for 48 hours until 40% CPE was observed. RNA was extracted by the addition of 1 ml of TRIzol reagent (ThermoFisher), processed for direct RNA sequencing using an Oxford Nanopore flow cell and the sequence reads were mapped to the SARS-CoV-2 genome as previously described (*14*).

### COVID-19 Genomics UK (COG-UK) Consortium Sequencing of Patient Isolates

Next generation sequencing of SARS-CoV-2 positive samples was performed by the COG-UK consortium (*15*) on nine samples taken from May, 2020 to February, 2021 (Tables 1 and 2). Sequencing at COG-UK sites was performed using a combination of Oxford Nanopore or Illumina platforms. Protocols used for each instrument were as described by COG-UK and indicated in Table 1. Library preparation and sequencing was performed for the majority of samples using the MinION platform (Oxford Nanopore, UK) with the nCoV-2019 sequencing protocol v3 (LoCost) (*16*). There was no protocol data available for the three samples sequenced on Illumina platforms.

**Table 1:** SARS-CoV-2 genome detection in swab samples.

| *Day of sample | Initial result | Initial Assay Used | Initial Ct/ RLU | <sup>a</sup>NMD Ct Value N | <sup>a</sup>NMD Ct Value NSP2 | Sample Type<sup>b</sup> | NGS Prot<sup>c</sup> | GISAID<sup>d</sup> | ENA<sup>e</sup> |
| --- | --- | --- | --- | --- | --- | --- | --- | --- | --- |
| D1 | + | Altona | E-23.98, S-24.51 | 19.25 | 20.87 | N&T | ILL-NS, ? | EPI_ISL_469397 | ERR4422451 |
| D58 | + | TMA | RLU-1087 |  |  | N&T | ILL-NS, ? |  | ERR4616078 |
| D94 | + | TMA | RLU-1193 | 18.11 | 19.50 | N&T |  |  |  |
| D124 | + | TMA | RLU-1256 |  |  | N&T |  |  |  |
| D147 | + | TMA | RLU-1190 |  |  | N&T |  |  |  |
| D155 | + | TMA | RLU-1291 | 19.69 | 20.30 | N&T | ILL-HS, ? |  | ERR4780220 |
| D164 | + | TMA | RLU-1275 |  |  | N&T |  |  |  |
| D180 | + | TMA | RLU-1297 |  |  | N&T |  |  |  |
| D209 | + | TMA | RLU-1172 | 19.19 | 19.19 | N&T |  |  |  |
| D217 | + | TMA | RLU-1181 | 18.79 | 20.23 | N&T |  |  |  |
| D222 | + | TMA | RLU-1173 | 20.48 | 22.06 | N&T | OxN, LC |  |  |
| D227 | + | TMA | RLU-1194 | 21.99 | 23.32 | N&T | OxN, LC |  |  |
| D230 | + | TMA | RLU-1238 | 19.12 | 20.29 | N&T |  |  |  |
| D263 | + | TMA | RLU-1354 | 24.19 | 24.77 | N&T | OxN, LC |  | ERR5510862 |
| D266 | + | TMA | RLU-1312 |  |  | N&T | OxN, LC | EPI_ISL_1103757 | ERR5405452 |
| D271 | + | TMA | RLU-1180 | 23.48 | 24.55 | N&T | OxN, LC |  | ERR5502292 |
| D276 | + | TMA | RLU-1303 | 31.60 | Neg | Nasal | QC-Fail |  |  |
| D283 | + | TMA | RLU-1284 | 33.28 | 33.41 | N&T | QC-Fail |  |  |
| D286 | + | TMA | RLU-1304 | 32.88 | 32.78 | Throat |  |  |  |
| D290 | + | TMA | RLU-1260 | 30.61 | 31.24 | Nasal | OxN, LC |  | ERR5498086 |
| D297 | +/- | TMA | RLU-491 | Neg | Neg | Nasal |  |  |  |
| D306 | + | TMA | RLU-1181 | 22.35 | 23.40 | Nasal |  |  |  |
| D311 | - | TMA | RLU-307 |  |  | N&T |  |  |  |
| D318 | - | TMA | RLU-287 |  |  | Nasal |  |  |  |
| D320 | - | TMA | RLU-309 | Neg | Neg | N&T |  |  |  |

\* Day of first positive RT-PCR test = day 1.
<sup>a</sup>NMD = Closed NeuMoDx SARS-CoV-2 RT-PCR commercial assay; <sup>b</sup>N&T = nose and throat
<sup>c</sup>NGS sequencing protocol. ILL-NS = Illumina NovaSeq , ILL-HS = Illumina HiSeq2500 , OxN = Oxford Nanopore MinION, ? = protocol unknown; LC = nCoV-2019 LoCost V3 (16).
<sup>d</sup>GISAID (www.gisaid.org/, (47)) sequence accession number.
<sup>e</sup>ENA (www.ebi.ac.uk/ena) European nucleotide archive accession number.

**Table 2:**
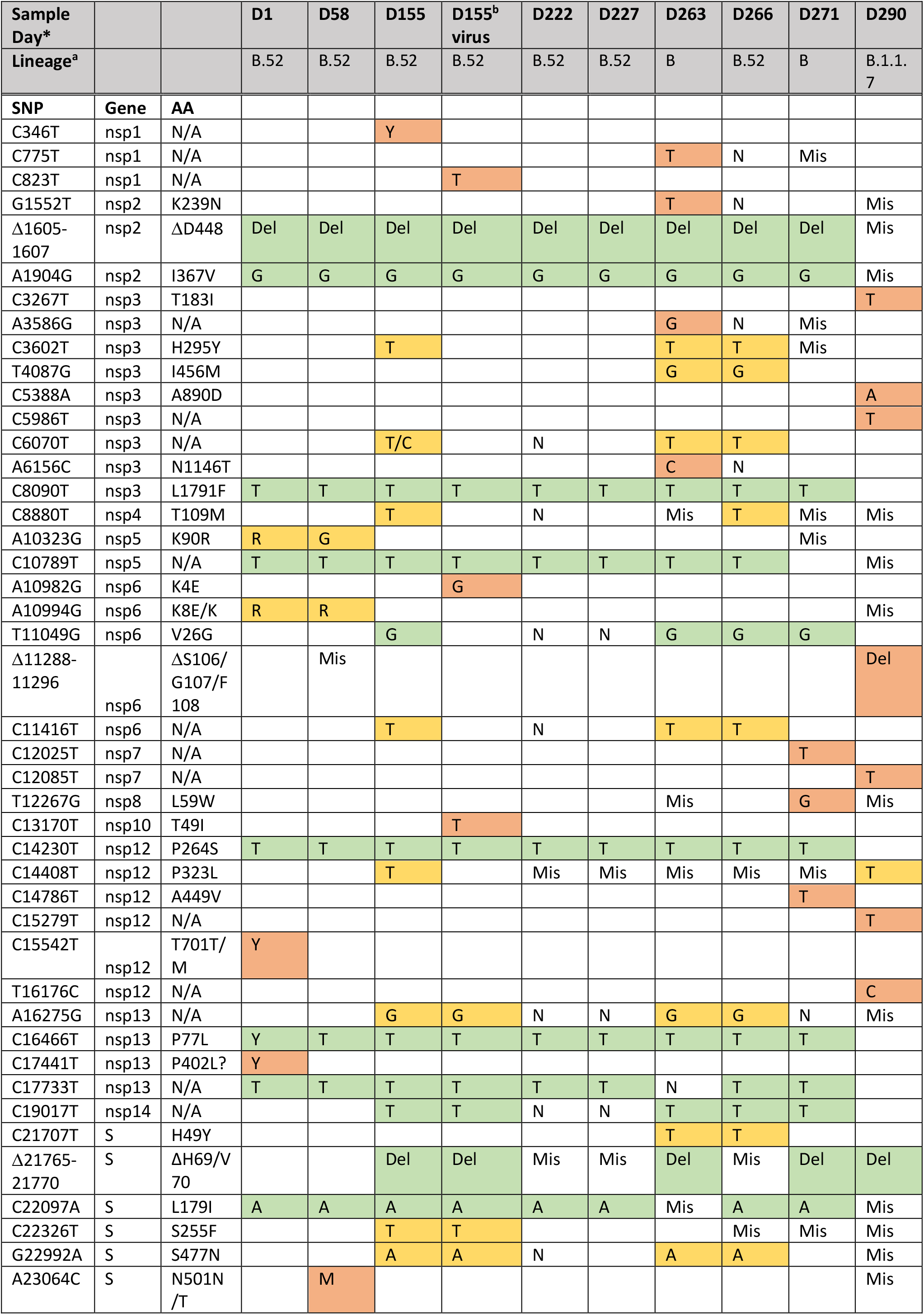
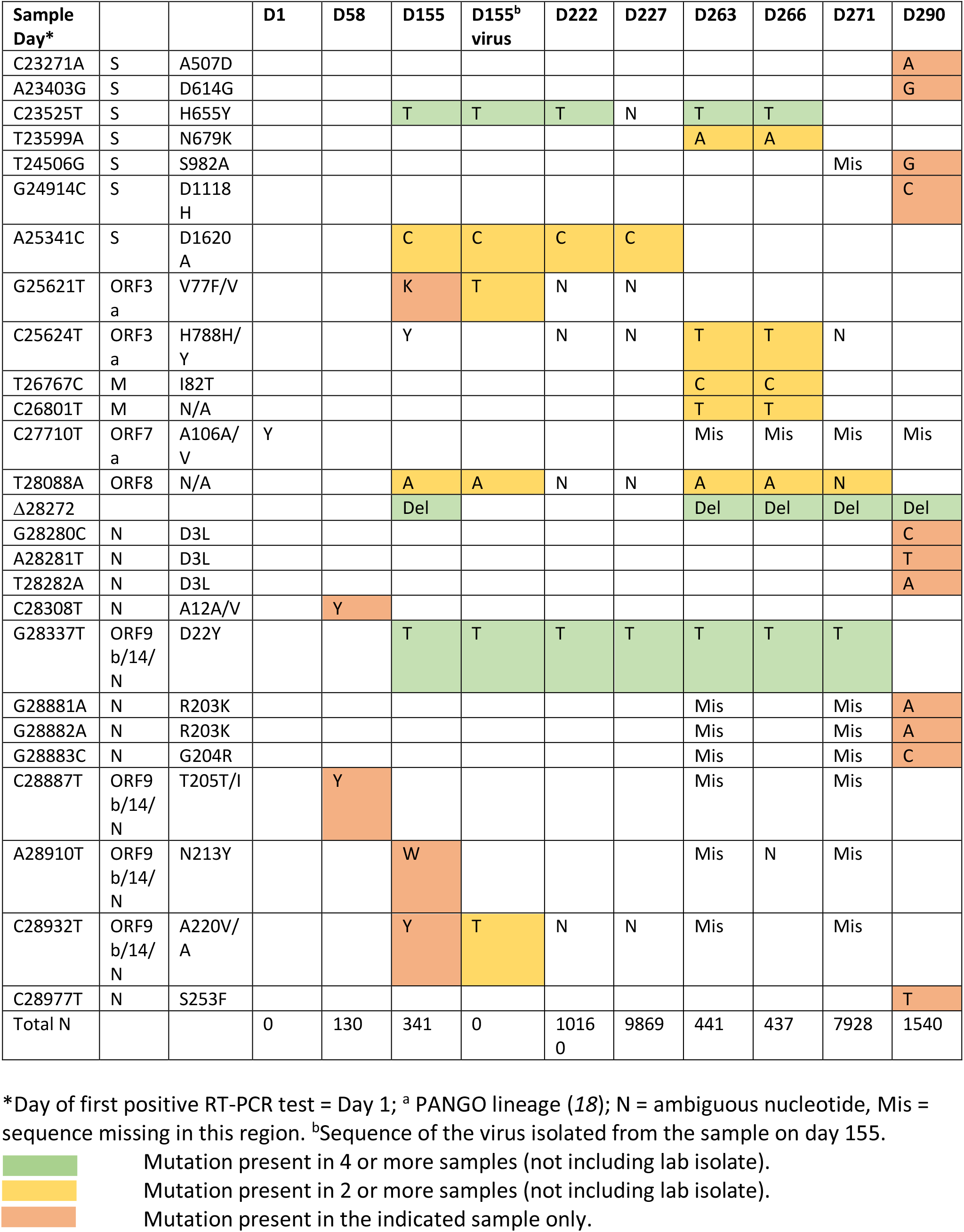
Summary of identified mutations compared to the SARS-CoV-2 Wuhan-Hu-1 sequence.

Consensus FASTA sequences were downloaded from the MRC-CLIMB database (*17*) and raw fastq files from the European Nucleotide Archive (ENA), or were provided directly from sequencing centres. Sequencing webtools including Pangolin (*18*), CoV-GLUE (*19*) and NextClade (*20*) were used to explore mutations and assign lineages. The raw fastq files were mapped to the SARS-CoV-2 reference genome (Wuhan-Hu-1; GenBank: MN908947.3) using MiniMap2-2.17 (with –ax and sr flags set) (*21*) and the mapped data was analysed with in house scripts to detect and quantify background insertions and deletions. In addition, mapped reads were manually inspected where appropriate to ascertain if dominant mutations at later time points were present at low levels in previous time points.

### Remdesivir sensitivity assay

VeroE6/TMPRSS2 cells were seeded for 18 hours in DMEM before being infected for 1 hour at 37 °C with hCoV-19/England/BRIS-MKW1/2020 or SARS-CoV-2/human/Liverpool/REMRQ0001/2020 at an MOI of 0.02 for 60 mins at 37 °C. The virus inoculum was then removed and MEM media containing remdesivir (starting concentration 20 µM) added in triplicate in a half-log 8-point dilution series. Cells were incubated at 37 °C for 4 days until cytopathic effects were visible. Sensitivity of the virus isolates to inhibition by remdesivir was assessed by MTT assay as previously described (*13*). Viable cells were calculated and the % inhibition of virus induced cell death was determined and plotted as a function of media and virus only controls. Graphs were plotted in GraphPad Prism v8.4.3.

### Casirivimab and imdevimab treatment

The hospital trust drug and therapeutics committee approved the off-licence use of casirivimab and imdevimab and the UK Medical and Healthcare products Regulatory Agency authorised importation. The therapy was supplied free of charge on a compassionate use basis by Regeneron Pharmaceuticals.

## Results

### Case description

A full description of the case history is available from the authors on request.

### Viral load during the course of a persistent SARS-CoV-2 infection

A male in his 8^th^ decade was admitted to hospital with respiratory symptoms and had a positive nasopharyngeal RT-PCR test for SARS-CoV-2 (Altona RealStar® SARS-CoV-2 RT-PCR; day 1 in Figure 1, Tables 1 and 2). He was neutropenic and lymphopaenic (neutrophils 0.9 × 10^6^ cells/L, lymphocytes 0.52 × 10^6^ cells/L). The patient had previously been treated for chronic lymphocytic leukaemia (CLL; six cycles of fludarabine/cyclophosphamide/rituximab) and was hypogammaglobulinaemic. He was randomised to the azithromycin arm of the RECOVERY study (*22*). The presence of viral RNA in swab samples obtained from the patient was monitored over the next 320 days using the Aptima SARS-CoV-2 transcription mediated assay (TMA), with quantitation of the viral load and sequencing of the viral genome done at specific time points by RT-PCR (NeuMoDx SARS-CoV-2 assay) and NGS respectively (Figure 1, Table 1). The patient was treated to eliminate SARS-CoV-2 and the associated COVID-19 symptoms with; remdesivir (a 17-day treatment), intravenous human immunoglobulin (IVIG; on day 4 of the remdesivir treatment) and finally casirivimab and imdevimab, a combination of two synthetic monoclonal antibodies targeted at different SARS-CoV-2 spike protein epitopes (*23, 24*). A time course of the patient’s admissions, TMA/RT-PCR results and treatment times are shown in Figure 1 and Table 1. Quantitation of viral RNA over the 320-day period indicated stable, high levels of viral RNA throughout the clinical course (ranging from C_t_ values of 18.11 - 24.19 and 19.19 - 24.55 for the N and nsp2 target genes respectively; Table 1) prior to casirivimab/imdevimab treatment. Nasopharyngeal swab tests then remained RT-PCR positive for SARS-CoV-2 for 41 days post-treatment (days 266-306; with one indeterminate on day 33). The first negative result was taken 46 days after treatment (day 311) with all subsequent samples remaining negative (up to day 114 post-treatment, at the time of writing).

**Figure 1:**
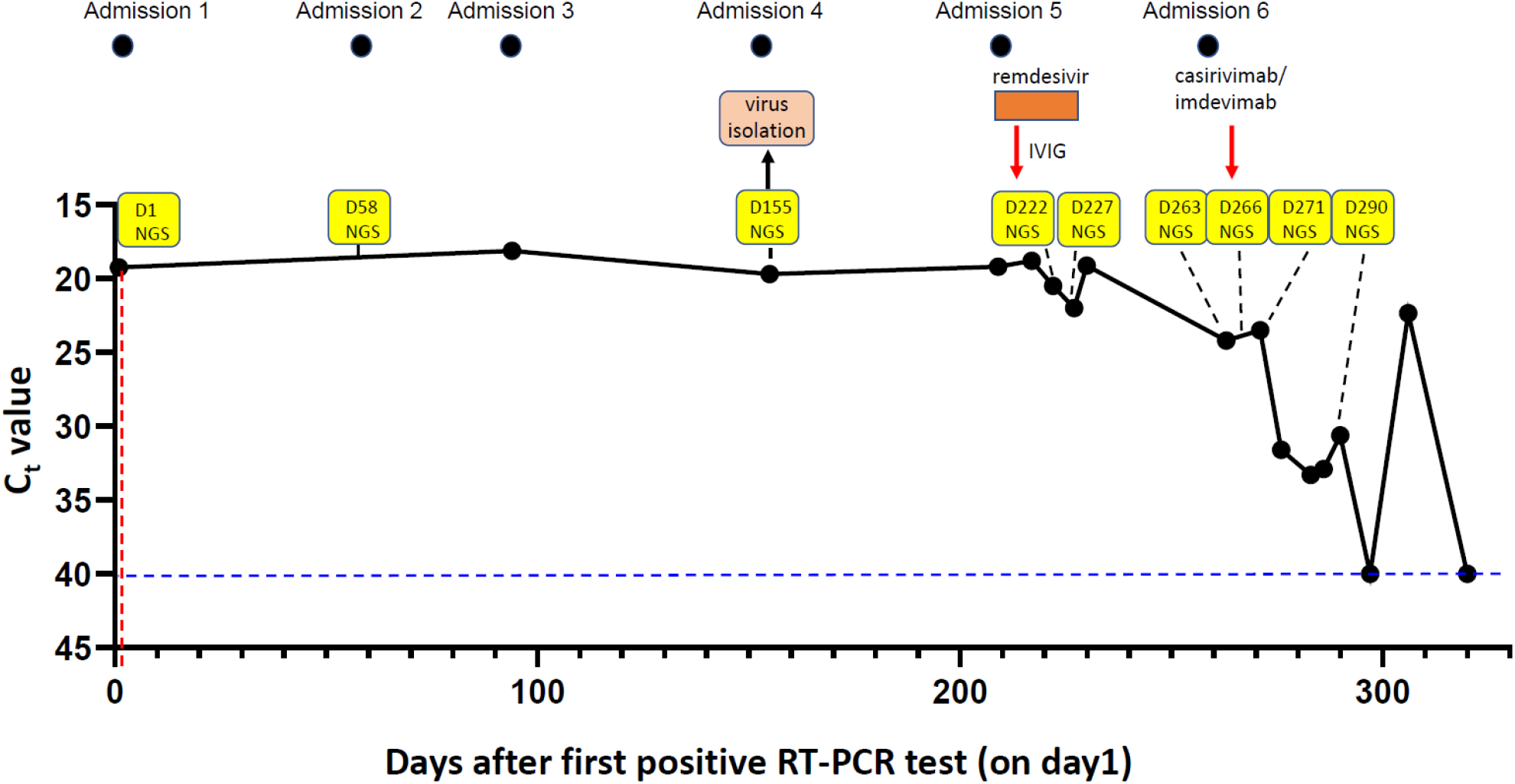
Clinical and sampling timeline. Patient swab samples were collected and tested for SARS-CoV-2 by RT-PCR. The results of the NeuMoDx RT-PCR assay are plotted as the Ct value for the N gene target over the time course of infection with the first positive RT-PCR test done on “day1 (D1)”. All Ct values above the horizontal blue dashed line showed a positive result. The times of hospital admission, anti-SARS-CoV-2 treatments, virus isolation and sample collection for NGS are indicated.

### Genomic sequencing

Next generation sequence (NGS) analysis of SARS-CoV-2 positive samples was performed by the COG-UK consortium (*15*) on nine samples taken from day 1 to day 290 (Figure 1, Tables 1 and 2). Of these, three samples resulted in poor genome sequence coverage (days 222, 227 and 290) but were included in the analysis as they could be used to confirm the virus lineage. The viral lineage for all samples was identified as B.52 based on PANGO classification (pangolin.cog-uk.io (*18*)), except for the day 271 and 290 samples (albeit with poorer sequence coverage) defined as lineage B and B.1.1.7 respectively. The B.52 lineage was first detected in the UK in March 2020 (*18*). Lineage defining mutations which were found in all genomes analysed prior to casirivimab/imdevimab treatment included; spike L179I, nsp2 ΔD268, nsp2 I367V, nsp3 L1791F, nsp12 P264S and nsp13 P77L (Table 2). Although the viral sequence on day 155 was defined as lineage B.52, it had undergone rapid evolution compared to the sequences obtained from the day 1 and 58 samples. A number of additional important mutations were identified in this and the viral genome sequences determined from subsequent samples. Significantly, there were four non-synonymous nucleotide substitutions in the spike gene resulting in the mutations; S255F, S477N, H655Y and D1620A, in addition to deletion of nucleotides 21765-21770, resulting in deletion of amino acids H69 and V70 (ΔH69/ΔV70). Except for S255F, the new spike protein mutations were identified in subsequent viral genome sequences except for the genome sequence obtained from the day 290 sample (with the exception of ΔH69/ΔV70). The mutations ΔH69/ΔV70 and H655Y have been identified in the SARS-CoV-2 variants of concern (VOCs) B.1.1.7 and P1 respectively (*25, 26*) whilst residue S477 is in the spike receptor binding domain (RBD) with the S477N substitution commonly found in other lineages (*2*). The mutation S477N has previously been detected during persistent infection of an immunocompromised patient (*27*). More detailed analysis of low frequency variants in the viral genome sequences revealed that in the sequence from the day 58 sample there was already evidence of the emergence of the ΔH69/ΔV70 and H655Y mutations with 4.7 and 19.3% of the reads specifying these mutations respectively (Table 3). Moreover, in the first sample taken in May 2020 (day 1) some 0.5% of the sequence reads in that region already had the ΔH69/ΔV70 deletion. By contrast, the mutation S477N only became detectable in the day 155 sample. In addition, at nucleotide 23064 within the S gene, there was a mixed nucleotide population in the sequence from the day 58 sample with 69% of the nucleotides changed to a C from an A. Thus, this sample had a mixed sequence population specifying the wild type N501 and a N501T mutation whereas in the other samples this position was >95% wild type. The change N501Y is present in the three SARS-CoV-2 VOCs B.1.1.7, B.1.351(501Y.V2) and P1 (*25, 26, 28*). Further non-synonymous substitutions were identified in the day 155 and subsequent samples, outside of the spike gene, resulting in the mutations; nsp3 H295Y, nsp4 T109M, nsp6 V26G and nsp12 P323L, the latter was only found in the viral sequence from the day 155 sample (Table 2).

**Table 3:** Low frequency variant analysis of the spike protein mutations S477N, N501T, H655Y and the ΔH69/ΔV70 deletion.

| Mutation | Day | Total* Reads | G (%)# | A (%) | T (%) | C (%) | Consensus |
| --- | --- | --- | --- | --- | --- | --- | --- |
| **S477N**<br>**(G22992→A)** | **1** | 86194 | 86025 (99.8) | 26 (0.000) | 98 (0.1) | 45 (0.1) | G |
|  | **58** | 19404 | 19384 (99.9) | 1 (0.01) | 16 (0.08) | 3 (0.02) | G |
|  | **155** | 4098 | 273 (6.7) | 3825 (93.3) | 0 | 0 | A |
| **N501T**<br>**(A23064→C)** | **1** | 46638 | 4 (0) | 44448 (95.3) | 72 (0.2) | 2113 (4.5) | A |
|  | **58** | 10443 | 2 (0) | 3214 (30.8) | 9 (0.1) | 7218 (69.1) | C |
|  | **155** | 3756 | 0 (0) | 3753 (99.9) | 3 (0.1) | 0 (0) | A |
| **H655Y**<br>**(C23525→T)** | **1** | 0 (0) | 11 (0.1) | 57 (0.3) | 16910 (99.6) | 0 (0) | C |
|  | **58** | 0 (0) | 4 (0) | 1726 (19.3) | 7219 (80.7) | 0 (0) | C |
|  | **155** | 2 (0) | 1 (0) | 6845 (92.3) | 565 (7.6) | 2 (0) | T |
|  |  |  | **H69/V70 count (%)** | **ΔH69/ΔV70 count (%)** |  |  |  |
| **ΔH69/ΔV70**<br>**(Δ21765-21770)** | **1** | 24942 | 24813 (99.5) | 129 (0.5) |  |  | TACATG |
|  | **58** | 8459 | 8061 (95.3) | 398 (4.7) |  |  | TACATG |
| | **155** | 4987 | 390 (7.8) | 4597 (92.2) | | | ΔTACATGΔ |

\* Total read counts G, A, T and C.
#Count for each nucleotide (for G, A, T and C) and % of the total reads.

Six RT-PCR positive samples (days 222-290, Table 1) collected during and post-remdesivir treatment of the patient were used for sequencing. The patient was also administered IVIG four days after the start of remdesivir treatment (Figure 1). During the treatment, no sequence changes occurred relative to the last viral genome sequence prior to treatment (day 155), albeit the sequence coverage was poor using the day 222 and 227 samples, taken 9 and 14 days after the remdesivir treatment commenced. A number of new nucleotide substitutions were detected in the viral genome sequences sampled approximately one month after the end of treatment (days 263 and 266), including three synonymous and six non-synonymous changes (in the nsp2, nsp3, S and M genes, Table 2). There were a further two synonymous and two non-synonymous changes that had been observed in earlier genome sequences.

The treatment with casirivimab/imdevimab commenced on day 265. On day 263, the viral swab showed a C_t_ value of 24.2/24.6 (N and nsp2 genes respectively) and did not change significantly by day 271. As described above, the genome sequence obtained from the day 271 sample did not show evolution compared to the previous genome sequences (Table 2). Although the patient swabs were positive for SARS-CoV-2 over the next 34 days, the viral load decreased significantly with C_t_ values > 30, with the exception of the day 306 sample which decreased to 23.4/24.4. The patient samples were then negative for the presence of SARS-CoV-2 RNA. Interestingly, sequencing of RNA from the day 290 sample suggested that the virus lineage was B.1.1.7, which was the predominant circulating lineage. There was no evidence that the patient was co-infected with the B.52 lineage, suggesting the possibility that this virus was cleared and that the patient had been transiently re-infected with the B.1.1.7 virus before total clearance of the virus.

### Viral culture and remdesivir susceptibility

Given the repeated admissions and ongoing RT-PCR positivity, the sample from day 155 was used for viral isolation to establish whether infectious SARS-CoV-2 could be recovered from the patient. An aliquot of the nasopharyngeal swab sample that was confirmed SARS-CoV-2 positive by RT-PCR (C_t_ value = 19.7/20.3 for the N/nsp2 genes respectively) was passaged on VeroE6/TMPRSS2 cells for 13 days until consistent CPE was visible. Supernatant from these cells was filtered and qRT-PCR analysis returned a C_t_ value of 11.25. To confirm the presence of replicating virus in the supernatant, it was titrated by a 2-fold dilution series and inoculated onto cells. Cells were subjected to immunofluorescence analysis and shown to be positive for the presence of the SARS-CoV-2 N protein and intracellular dsRNA (Figure 2A and B). This confirmed the presence of infectious virus and ongoing chronic patient infection for at least 6 months after symptom onset. Analysis of the growth kinetics of the isolated virus compared to SARS-CoV-2/human/Liverpool/REMRQ0001/2020 isolated in May, 2020 and the hCoV-19/England/204690005/2020, a B.1.1.7 lineage virus, showed they had comparable growth kinetics in VeroE6/TMPRSS2 and human Caco-2-ACE2 cells over the period 1 – 3 days post infection (Figure 2C). NGS analysis of the isolated virus showed that it had acquired one synonymous and two non-synonymous mutations (nsp6 K4E and nsp10 T49I) relative to the sequence obtained from the clinical sample on day 155 from which it was derived but retained all the other B.52 lineage defining mutations and the spike gene mutations ΔH69/ΔV70, S477N, H655Y and D1620A (Table 2).

**Figure 2:**
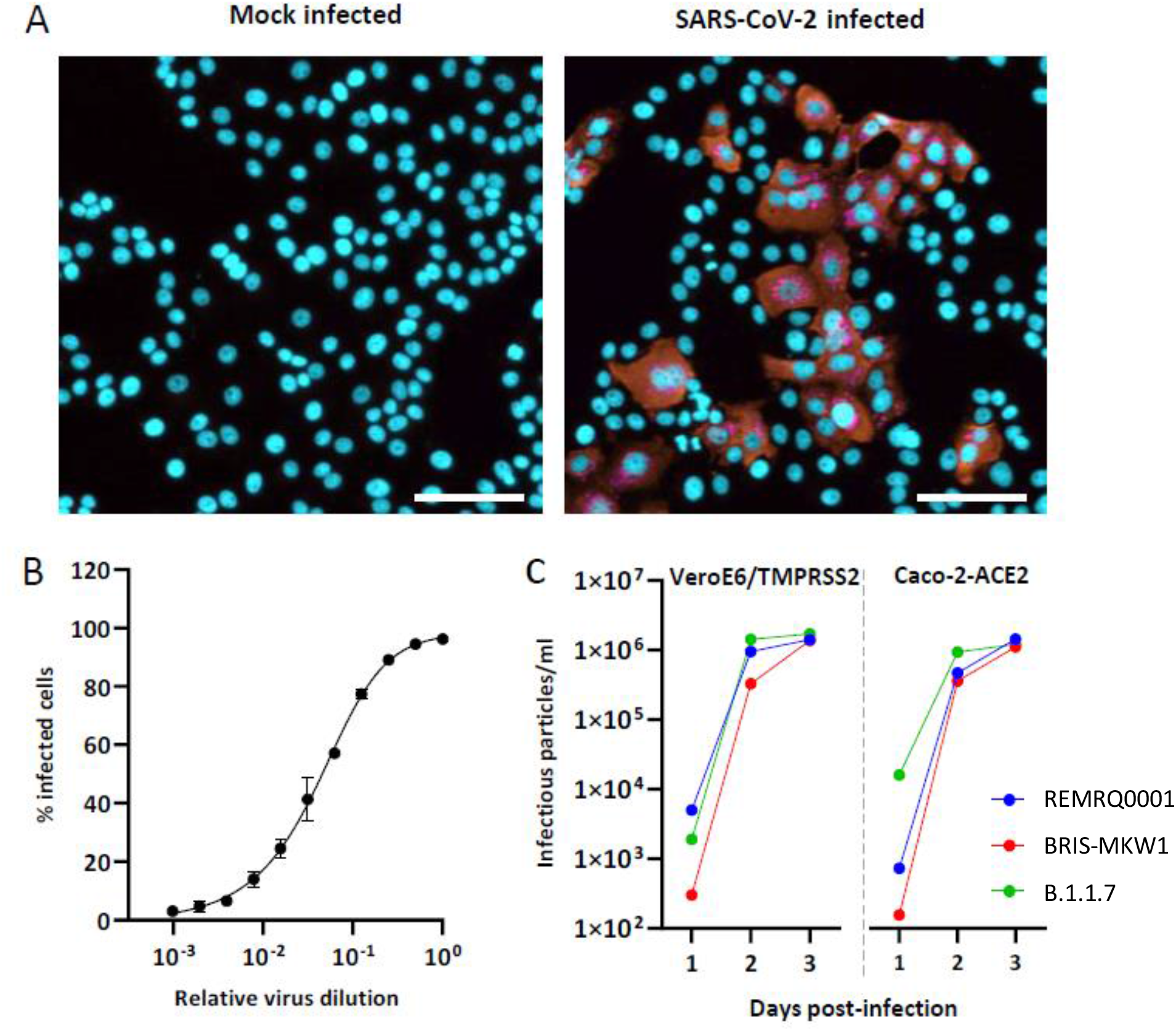
Confirmation of infectious virus isolation from the immunocompromised patient. **A)** VeroE6/TMPRSS2 cells were mock infected or infected with the culture supernatant from cells showing CPE after inoculation with an aliquot of the day 155 clinical sample. After 18 hours the cells were fixed, permeabilised and stained for DAPI (blue), SARS-CoV-2 N protein (orange) and dsRNA (pink). **B)** A stock of the clinical virus isolate was titrated on VeroE6/TMPRSS2 cells and the % of infected cells determined by detection of SARS-CoV-2 N protein associated with nuclei. Image acquisition, analysis and quantitation were performed on the ImageXpress Pico imaging system. Scale bar shows 100 μm. **C)** The growth kinetics of the isolated virus (BRIS-MKW1) was compared those of SARS-CoV-2/human/Liverpool/REMRQ0001/2020 (REMRQ0001) and hCoV-19/England/204690005/2020 (B.1.1.7) on VeroE6/TMPRSS2 and Caco-2-ACE2 cells. The cells were infected at an MOI of 0.01 and the amount of viral RNA present in the culture supernatants determined at 1, 2 and 3 days post-infection by qRT-PCR. The amount of infectious particles/ml was determined by comparison to a calibrated standard.

To test the hypothesis that the patient’s failure to respond to remdesivir treatment might have resulted from the acquisition of viral resistance, a drug sensitivity assay was performed. The IC_50_ values for remdesivir were 1.4 and 2.4 µM for SARS-CoV-2/human/Liverpool/REMRQ0001/2020 and hCoV-19/England/BRIS-MKW1/2020 respectively, which were not significantly different (Figure 3), suggesting that the failure to produce a change in viral load in the patient or any improvement in clinical state was not due to pre-existing mutations conferring remdesivir resistance.

**Figure 3:**
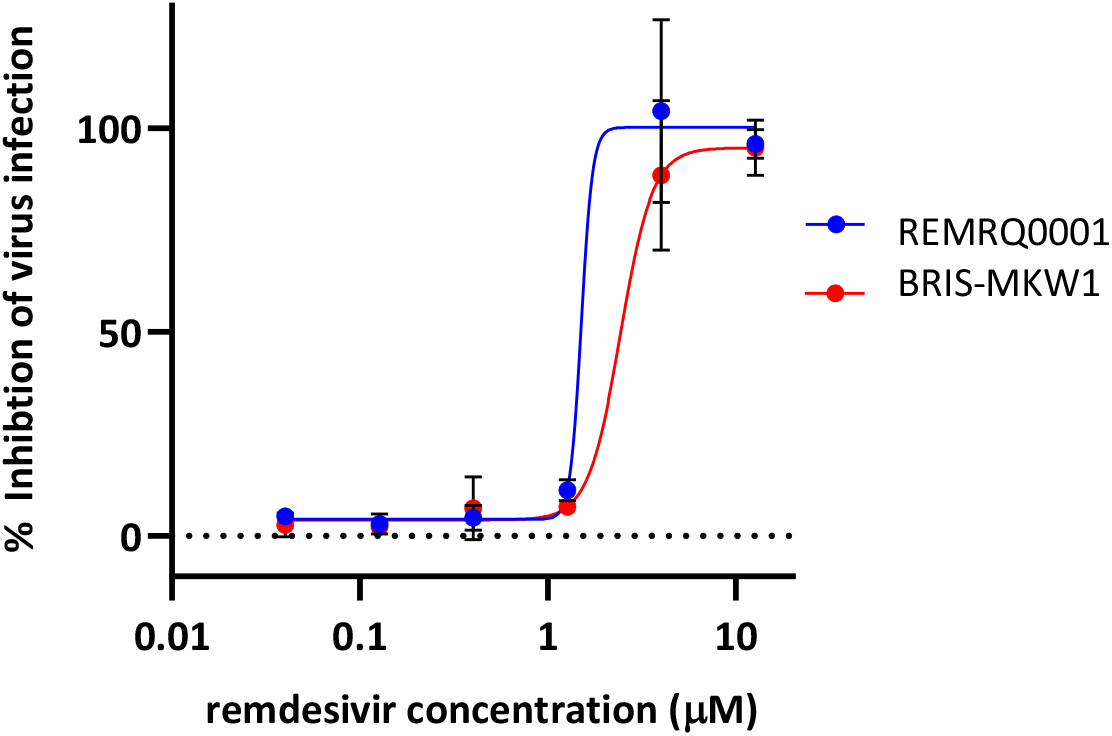
Remdesivir sensitivity of the SARS-CoV-2 clinical isolate. VeroE6/TMPRSS2 cells were infected with hCoV-19/England/BRIS-MKW1/2020 (BRIS-MKW1, red) or SARS-CoV-2/human/Liverpool/REMRQ0001/2020 (REMRQ0001, blue) at an MOI of 0.02, for 1 hour. The inoculum was removed and replaced with media containing a dilution series of remdesivir (12.5 µM starting concentration) and the cells incubated for 3 days at 37 °C. The % inhibition of virus induced cell death was determined by MTT assay and plotted as a function of media and virus only controls. Error bars show the variance of triplicate wells. The IC50 values were 1.4 and 2.4 µM for SARS-CoV-2/human/Liverpool/REMRQ0001/2020 and hCoV-19/England/BRIS-MKW1/2020 respectively.

## Discussion

To our knowledge, we report the most protracted, viral culture confirmed, case of SARS-CoV-2 infection. Viral culture was achieved from a sample obtained 155 days after the first RT-PCR positive test, and 197 days after symptom onset. Active viral infection was further confirmed by the successive acquisition of certain single-nucleotide polymorphisms (SNPs) and deletions in viral sequences obtained from nasopharyngeal samples taken over the course of his illness. These mutations arose spontaneously in the absence of immunotherapy or antiviral drug treatment with one, the ΔH69/ΔV70 deletion, previously described and attributed to the treatment of an individual persistently infected with SARS-CoV-2 with convalescent plasma (*10*). Given the hypogammaglobulinaemic state of the patient in this study and the absence of specific SARS-CoV-2 antibody on testing, viral evolution in this case was potentially driven by T-cell selection and resultant viral fitness.

After five months of persistent SARS-CoV-2 replication in this immunocompromised patient, the virus was found to have evolved rapidly between the day 58 and 158 samples. Specifically, it acquired six additional non-synonymous SNPs and a deletion which then became fixed in the viral genome over the duration of infection. There were three specific mutations in the spike protein for which there is evidence of biological effect. The ΔH69/ΔV70 deletion is present in the B.1.1.7 VOC (*26*). The mutation came to attention as one of a cluster of five mutations in the spike protein associated with spill over of SARS-CoV-2 to mink. Of the five mutations, a combination of ΔH69/ΔV70 and Y453F are sufficient to evade neutralising antibody recognition (*29*). The same combination was identified in the viral sequence present in a persistently infected immunocompromised patient (*30*), suggesting convergent evolution. The emergence of the ΔH69/ΔV70 mutation in an immunocompromised individual after convalescent plasma therapy was accompanied by the spike mutation D796H, suggesting a synergistic effect which mediated antibody escape (*10*). More detailed characterisation of the effects of the ΔH69/ΔV70 mutation, either alone or in combination with other spike mutations, demonstrated that ΔH69/ΔV70 did not lead to increased susceptibility to serum neutralisation but rather enhanced SARS-CoV-2 infectivity by altering spike processing, potentially compensating for RBD mutations that decrease infectivity but lead to antibody escape (*31*). The results of this study, in which the ΔH69/ΔV70 mutation was acquired in the absence of antibody pressure support this idea. The two spike protein mutations K477N and H655Y accompanied the identification of the ΔH69/ΔV70 deletion in the virus sequence determined from the day 155 sample. The residue S477 is located in the spike protein RBD with S477N the fourth most frequent RBD substitution identified in clinical isolates (*2*). In a recent study, the S477N mutation was found to confer resistance to neutralisation by a broad range of mAbs but not convalescent serum (*32*) and was found to occur in a persistently infected patient treated with CP (*27*). The mutation H655Y has been shown to mediate escape from mAb neutralisation using a viral pseudotype system (*33*) and was identified to occur during transmission in hamsters (*34*) and cats (*35*). The combination of the acquired mutations ΔH69/ΔV70, S477N and H655Y may therefore have altered the infectivity of SARS-CoV-2 and played a role in maintaining the persistent infection. Low frequency variant analysis of the mutations suggested the S477N mutation only arose after emergence of the ΔH69/ΔV70 and H655Y mutations. Interestingly, all three mutations were maintained in the virus isolated from the clinical sample.

Attempts to clear or even reduce viral load with an extended course of remdesivir were not successful. The reason for this is not clear as *in vitro* sensitivity was confirmed. A number of additional mutations were identified in the viral genome after remdesivir treatment, but they did not correspond with mutations known to confer resistance to remdesivir *in vitro* for either SARS-CoV-2 (nsp6 I168T and nsp12 E208D (*36*) or other betacoronaviruses (*37*). The use of remdesivir to clear persistently replicating SARS-CoV-2 from other immunocompromised patients has typically resulted in an initial suppression of virus replication followed by relapse after cessation of treatment (*10, 38-40*). Collectively the results suggest that remdesivir treatment needs to be prolonged or optimally combined with other treatments to eradicate SARS-CoV-2 from immunocompromised individuals. In the absence of an effective humoral response antivirals may have limited benefit.

There have been relatively few published descriptions of persistent SARS-CoV-2 infection, the majority associated with B cell immunodeficiency (*7, 10, 27, 41-44*). Conversion to PCR negativity and dramatic symptomatic improvement was achieved only after administration of casirivimab and imdevimab. This synthetic IgG1 neutralising antibody cocktail targets two epitopes on the viral spike protein with the aim of reducing viral load and preventing the rapid mutational escape observed with single antibody therapy (*23, 24*). Studies in outpatient cohorts with early COVID-19 have shown it to be safe and associated with reductions in viral load, particularly among those who were SARS-CoV-2 serum-antibody negative at baseline (*45*). At present they are authorised in the USA and EU for use in non-hospitalised patients with mild to moderate disease. Trials among acutely unwell hospitalised patients with more severe disease manifestations are ongoing.

Treating this immunocompromised patient group is a challenge. Where antivirals have been used they may ease symptoms and have been associated with sustained improvement in some patients, yet appear to produce only a transient drop in viral load with recrudescence once treatment stops in others. Antibody therapy, whether with CP or synthetic monoclonal cocktails holds promise. There are reports of improvement and complete symptom resolution with viral clearance in patients with sustained illness with both forms of antibody treatment (*8, 9, 41, 42*). Yet even as this form of immunotherapy finds a role in prevention or treatment of very early infection (*46*) it is falling out of favour as a treatment for acutely unwell immunocompetent patients hospitalised with COVID-19 (*22*), given the negative results of the large RECOVERY trial with convalescent plasma. However, it is likely that those with chronic persistent infection will respond differently. No randomised controlled trial conducted specifically within this patient group has yet been reported. It is important that any potential benefit for this small but important cohort is not overlooked. Beyond treatment of sick individuals there are compelling public health arguments for attempting cure. Analysis of viral sequences obtained from such patients over time suggests, as with the case described here, an accelerated rate of viral evolution within immunosuppressed individuals potentially facilitating the emergence of new strains within the wider population (*1, 7, 39*). Such treatments themselves risk driving the selection of resistance as suggested by others (*10*) thus any experimental treatment regime would require close monitoring of viral evolution and early recognition of any unintended consequences.

In conclusion, immunosuppressed patients, specifically those with humoral deficiencies, are vulnerable to chronic persistent SARS-CoV-2 infection. This may not result in acute illness necessitating prolonged hospitalisation, but rather lead to fatigue, malaise and slowly progressive debilitation with declining respiratory function. This potentially presents a unique infection control risk: individuals infected by virus acquiring new mutations who are, to a certain extent, active in the community. Whilst such cases are likely to be very rare, studies of potential therapies are urgently required, motivated by both compassion for the suffering of the individual, and the preservation of vaccine efficacy and hence the health of the population.

## Data Availability

All sequence data referred to in the manuscript is available through GISAID or the European Nucleotide Archive database.
www.gisaid.org
www.ebi.ac.uk/ena

## Funding and Acknowledgments

The DISCOVER study was funded by the Southmead Hospital Charity. Immunological work was funded by an Elizabeth Blackwell Institute TRACK award. A.D.D. / D.A.M are supported by the United States Food and Drug Administration (HHSF223201510104C) and the UK Research and Innovation / Medical Research Council (MRC) and Biotechnology and Biological Sciences Research Council (grants MR/V027506/1 and BB/V013874/1). M.K.W is supported by MRC grant MR/V027506/1 (awarded to A.D.D). COG-UK is supported by funding from the MRC part of UKRI, the National Institute of Health Research (NIHR) and Genome Research Limited, operating as the Wellcome Sanger Institute. The authors would like to acknowledge support of the University of Bristol’s Alumni and Friends, which funded the ImageXpress Pico Imaging System. A.D.D and D.A.M. are members of the G2P-UK National Virology consortium funded by MRC/UKRI (grant MR/W005611/1.) We acknowledge Professor Wendy Barclay (Imperial College, London, UK) and Professor Maria Zambon (Public Health England, UK) for kindly providing the SARS-CoV-2 B.1.1.7 isolate.

## Ethics

The DISCOVER study was ethically approved via the South Yorks REC (ref: 20/YH/0121, CRN approval no: 45469). The patient provided written informed consent.

