## Supplemental file 1_Appendix for "Chronic SARS-CoV-2 infection and viral evolution in a hypogammaglobulinaemic individual"

**Funding acquisition, Leadership and supervision, Metadata curation, Project administration, Samples and logistics, Sequencing and analysis, Software and analysis tools, and Visualisation:**
Dr Samuel C Robson PhD [13].

**Funding acquisition, Leadership and supervision, Metadata curation, Project administration, Samples and logistics, Sequencing and analysis, and Software and analysis tools:**
Prof Nicholas J Loman PhD [41] and Dr Thomas R Connor PhD [10, 69].

**Leadership and supervision, Metadata curation, Project administration, Samples and logistics, Sequencing and analysis, Software and analysis tools, and Visualisation:**
Dr Tanya Golubchik PhD [5].

**Funding acquisition, Metadata curation, Samples and logistics, Sequencing and analysis, Software and analysis tools, and Visualisation:**
Dr Rocio T Martinez Nunez PhD [42].

**Funding acquisition, Leadership and supervision, Metadata curation, Project administration, and Samples and logistics:**
Dr Catherine Ludden PhD [88].

**Funding acquisition, Leadership and supervision, Metadata curation, Samples and logistics, and Sequencing and analysis:**
Dr Sally Corden PhD [69].

**Funding acquisition, Leadership and supervision, Project administration, Samples and logistics, and Sequencing and analysis:**
Ian Johnston [99] and Dr David Bonsall PhD [5].

**Funding acquisition, Leadership and supervision, Sequencing and analysis, Software and analysis tools, and Visualisation:**
Prof Colin P Smith PhD [87] and Dr Ali R Awan PhD [28].

**Funding acquisition, Samples and logistics, Sequencing and analysis, Software and analysis tools, and Visualisation:**
Dr Giselda Bucca PhD [87].

**Leadership and supervision, Metadata curation, Project administration, Samples and logistics, and Sequencing and analysis:**
Dr M. Estee Torok FRCP [22, 101].

**Leadership and supervision, Metadata curation, Project administration, Samples and logistics, and Visualisation:**
Dr Kordo Saeed MD/ FRCPath [81, 110] and Dr Jacqui A Prieto PhD [83, 109].

**Leadership and supervision, Metadata curation, Project administration, Sequencing and analysis, and Software and analysis tools:**
Dr David K Jackson PhD [99].

**Metadata curation, Project administration, Samples and logistics, Sequencing and analysis, and Software and analysis tools:**
Dr William L Hamilton PhD [22].

**Metadata curation, Project administration, Samples and logistics, Sequencing and analysis, and Visualisation:**
Dr Luke B Snell MSc/ MBBS [11].

**Funding acquisition, Leadership and supervision, Metadata curation, and Samples and logistics:**
Dr Catherine Moore [69].

**Funding acquisition, Leadership and supervision, Project administration, and Samples and logistics:**
Dr Ewan M Harrison PhD [99, 88].

**Leadership and supervision, Metadata curation, Project administration, and Samples and logistics:**
Dr Sonia Goncalves PhD [99].

**Leadership and supervision, Metadata curation, Samples and logistics, and Sequencing and analysis:**
Prof Ian G Goodfellow PhD [24], Dr Derek J Fairley PhD [3, 72], Prof Matthew W Loose PhD [18] and Joanne Watkins MSc [69].

**Leadership and supervision, Metadata curation, Samples and logistics, and Software and analysis tools:**
Rich Livett MSc [99].

**Leadership and supervision, Metadata curation, Samples and logistics, and Visualisation:**
Dr Samuel Moses MD [25, 106].

**Leadership and supervision, Metadata curation, Sequencing and analysis, and Software and analysis tools:**
Dr Roberto Amato PhD [99], Dr Sam Nicholls PhD [41] and Dr Matthew Bull PhD [69].

**Leadership and supervision, Project administration, Samples and logistics, and Sequencing and analysis:**
Prof Darren L Smith PhD [37, 58, 105].

**Leadership and supervision, Sequencing and analysis, Software and analysis tools, and Visualisation:**
Dr Jeff Barrett PhD [99] and Prof David M Aanensen PhD [14, 114].

**Metadata curation, Project administration, Samples and logistics, and Sequencing and analysis:**
Dr Martin D Curran PhD [65], Dr Surendra Parmar PhD [65], Dr Dinesh Aggarwal MRCP [95, 99, 64] and Dr James G Shepherd MBChB/MRCP [48].

**Metadata curation, Project administration, Sequencing and analysis, and Software and analysis tools:**
Dr Matthew D Parker PhD [93].

**Metadata curation, Samples and logistics, Sequencing and analysis, and Visualisation:**
Dr Sharon Glaysher PhD [61].

**Metadata curation, Sequencing and analysis, Software and analysis tools, and Visualisation:**
Dr Matthew Bashton PhD [37, 58], Dr Anthony P Underwood PhD [14, 114], Dr Nicole Pacchiarini PhD [69] and Dr Katie F Loveson PhD [77].

**Project administration, Sequencing and analysis, Software and analysis tools, and Visualisation:**
Dr Alessandro M Carabelli PhD [88].

**Funding acquisition, Leadership and supervision, and Metadata curation:**
Dr Kate E Templeton PhD [53, 90].

**Funding acquisition, Leadership and supervision, and Project administration:**
Dr Cordelia F Langford PhD [99], John Sillitoe BEng [99], Dr Thushan I de Silva PhD [93] and Dr Dennis Wang PhD [93].

**Funding acquisition, Leadership and supervision, and Sequencing and analysis:**
Prof Dominic Kwiatkowski [99, 107], Prof Andrew Rambaut DPhil [90], Dr Justin O'Grady PhD [70, 89] and Dr Simon Cottrell PhD [69].

**Leadership and supervision, Metadata curation, and Sequencing and analysis:**
Prof Matthew T.G. Holden PhD [68] and Prof Emma C Thomson PhD/FRCP [48].

**Leadership and supervision, Project administration, and Samples and logistics:**
Dr Husam Osman PhD [64, 36], Dr Monique Andersson PhD [59], Prof Anoop J Chauhan [61] and Dr Mohammed O Hassan-Ibrahim PhD/FRCPath [6].

**Leadership and supervision, Project administration, and Sequencing and analysis:**
Dr Mara Lawniczak [99].

**Leadership and supervision, Samples and logistics, and Sequencing and analysis:**
Prof Ravi Kumar Gupta PhD [88, 113], Dr Alex Alderton PhD [99], Dr Meera Chand [66], Dr Chrystala Constantinidou PhD [94], Dr Meera Unnikrishnan PhD [94], Prof Alistair C Darby PhD [92], Prof Julian A Hiscox PhD [92] and Prof Steve Paterson PhD [92].

**Leadership and supervision, Sequencing and analysis, and Software and analysis tools:**
Dr Inigo Martincorena [99], Prof David L Robertson PhD [48], Dr Erik M Volz PhD [39], Dr Andrew J Page PhD [7] and Prof Oliver G Pybus DPhil [23].

**Leadership and supervision, Sequencing and analysis, and Visualisation:**
Dr Andrew R Bassett PhD [99].

**Metadata curation, Project administration, and Samples and logistics:**
Dr Cristina V Ariani PhD [99], Dr Michael H Spencer Chapman MBBS [99, 88], Dr Kathy K Li MBBCh/FRCPath [48], Dr Rajiv N Shah BMBS/MRCP/MSc [48], Dr Natasha G Jesudason MBChB MRCP FRCPath [48] and Dr Yusri Taha MD/PhD [50].

**Metadata curation, Project administration, and Sequencing and analysis:**
Martin P McHugh MSc [53], Dr Rebecca Dewar PhD [53].

**Metadata curation, Samples and logistics, and Sequencing and analysis:**

Dr Aminu S Jahun PhD [24], Dr Claire McMurray PhD [41], Ms Sarojini Pandey MSc [84], Dr James P McKenna PhD [3], Dr Andrew Nelson PhD [58, 105], Dr Gregory R Young PhD [37, 58], Dr Clare M McCann PhD [58, 105] and Mr Scott Elliott [61].

**Metadata curation, Samples and logistics, and Visualisation:**
Ms Hannah Lowe MSc [25].

**Metadata curation, Sequencing and analysis, and Software and analysis tools:**
Dr Ben Temperton Ph.D. [91], Dr Sunando Roy PhD [82], Dr Anna Price PhD [10], Dr Sara Rey PhD [69] and Mr Matthew Wyles [93].

**Metadata curation, Sequencing and analysis, and Visualisation:**
Stefan Rooke MSc [90] and Dr Sharif Shaaban PhD [68].

**Project administration, Samples and logistics, Sequencing and analysis:**
Dr Mariateresa de Cesare PhD [98].

**Project administration, Samples and logistics, and Software and analysis tools:**
Laura Letchford BSc [99].

**Project administration, Samples and logistics, and Visualisation:**
Miss Siona Silveira MSc [81], Dr Emanuela Pelosi FRCPath [81] and Dr Eleri Wilson-Davies MD/FRCPath [81].

**Samples and logistics, Sequencing and analysis, and Software and analysis tools:**
Dr Myra Hosmillo PhD [24].

**Sequencing and analysis, Software and analysis tools, and Visualisation:**
Áine O'Toole MSc [90], Dr Andrew R Hesketh PhD [87], Mr Richard Stark MSc [94], Dr Louis du Plessis PhD [23], Dr Chris Ruis PhD [88], Dr Helen Adams PhD [4] and Dr Yann Bourgeois PhD [76].

**Funding acquisition, and Leadership and supervision:**
Dr Stephen L Michell PhD [91], Prof Dimitris Grammatopoulos PhD/FRCPath [84, 112], Dr Jonathan Edgeworth PhD/FRCPath [12], Prof Judith Breuer MD [30, 82], Prof John A Todd PhD [98] and Dr Christophe Fraser PhD [5].

**Funding acquisition, and Project administration:**
Dr David Buck PhD [98] and Michaela John BSc [9].

**Leadership and supervision, and Metadata curation:**
Dr Gemma L Kay PhD [70].

**Leadership and supervision, and Project administration:**
Steve Palmer [99], Prof Sharon J Peacock [88, 64] and David Heyburn [69].

**Leadership and supervision, and Samples and logistics:**
Danni Weldon BSc [99], Dr Esther Robinson PhD [64, 36], Prof Alan McNally PhD [41, 86], Dr Peter Muir PhD [64], Dr Ian B Vipond PhD [64], Dr John BoYes MBChB [29], Dr Venkat Sivaprakasam PhD [46], Dr Tranprit Saluja FRCPath/MD [75], Dr Samir Dervisevic FRCPath [54] and Dr Emma J Meader FRCPath [54].

**Leadership and supervision, and Sequencing and analysis:**
Dr Naomi R Park PhD [99], Karen Oliver BSc [99], Dr Aaron R Jeffries Ph.D. [91], Dr Sascha Ott PhD [94], Dr Ana da Silva Filipe PhD [48], Dr David A Simpson PhD [72] and Dr Chris Williams MB BS [69].

**Leadership and supervision, and Visualisation:**
Dr Jane AH Masoli MBChB [73, 91].

**Metadata curation, and Samples and logistics:**
Dr Bridget A Knight PhD. [73, 91], Dr Christopher R Jones Ph.D. [73, 91], Mr Cherian Koshy MSc CSci FIBMS [1], Miss Amy Ash BSc [1], Dr Anna Casey PhD [71], Dr Andrew Bosworth PhD [64, 36], Dr Liz Ratcliffe PhD [71], Dr Li Xu-McCrae PhD [36], Miss Hannah M Pymont MSc [64], Ms Stephanie Hutchings [64], Dr Lisa Berry PhD [84], Ms Katie Jones MSc [84], Dr Fenella Halstead PhD [46], Mr Thomas Davis MSc [21], Dr Christopher Holmes PhD [16], Prof Miren Iturriza-Gomara PhD [92], Dr Anita O Lucaci PhD [92], Dr Paul Anthony Randell MBBCh [38, 104], Dr Alison Cox PhD [38, 104], Pinglawathee Madona [38, 104], Dr Kathryn Ann Harris PhD [30], Dr Julianne Rose Brown PhD [30], Dr Tabitha W Mahungu FRCPath [74], Dr Dianne Irish-Tavares FRCPath [74], Dr Tanzina Haque FRCPath PhD [74], Dr Jennifer Hart MRCP [74], Mr Eric Witele MSc [74], Mrs Melisa Louise Fenton DipHE [75], Mr Steven Liggett [79], Dr Clive Graham MD [56], Ms Emma Swindells Bsc [57], Ms Jennifer Collins BSc [50], Mr Gary Eltringham BSc [50], Ms Sharon Campbell MSc [17], Dr Patrick C McClure PhD [97], Dr Gemma Clark PhD [15], Dr Tim J Sloan PhD [60], Mr Carl Jones [15] and Dr Jessica Lynch PhD MBChB [2, 111].

**Metadata curation, and Sequencing and analysis:**
Dr Ben Warne MRCP [8], Steven Leonard PhD [99], Jillian Durham BSc [99], Dr Thomas Williams MD [90], Dr Sam T Haldenby PhD [92], Dr Nathaniel Storey PhD [30], Dr Nabil-Fareed Alikhan PhD [70], Dr Nadine Holmes PhD [18], Dr Christopher Moore PhD [18], Mr Matthew Carlile BSc [18], Malorie Perry MSc [69], Dr Noel Craine DPhil [69], Prof Ronan A Lyons MD [80], Miss Angela H Beckett MSc [13], Salman Goudarzi PhD [77], Christopher Fearn MRes [77], Kate Cook [77], Hannah Dent BSc [77] and Hannah Paul MRes [77].

**Metadata curation, and Software and analysis tools:**
Robert Davies [99].

**Project administration, and Samples and logistics:**
Beth Blane BSc [88], Sophia T Girgis MSc [88], Dr Mathew A Beale PhD [99], Katherine L Bellis [99, 88], Matthew J Dorman [99], Eleanor Drury [99], Leanne Kane [99], Sally Kay [99], Dr Samantha McGuigan [99], Dr Rachel Nelson PhD [99], Liam Prestwood [99], Dr Shavanthi Rajatileka PhD [99], Dr Rahul Batra MD [12], Dr Rachel J Williams PhD [82], Dr Mark Kristiansen PhD [82], Dr Angie Green PhD [98], Miss Anita Justice MSc [59], Dr Adhyana I.K Mahanama MD [81, 102] and Dr Buddhini Samaraweera MD [81, 102].

**Project administration, and Sequencing and analysis:**
Dr Nazreen F Hadjirin PhD [88] and Dr Joshua Quick PhD [41].

**Project administration, and Software and analysis tools:**
Mr Radoslaw Poplawski BSc [41].

**Samples and logistics, and Sequencing and analysis:**
Leanne M Kermack MSc [88], Nicola Reynolds PhD [7], Grant Hall BS [24], Yasmin Chaudhry BSc [24], Malte L Pinckert MPhil [24], Dr Iliana Georgana PhD [24], Dr Robin J Moll PhD [99], Dr Alicia Thornton [66], Dr Richard Myers [66], Dr Joanne Stockton PhD [41], Miss Charlotte A Williams BSc [82], Dr Wen C Yew PhD [58], Alexander J Trotter MRes [70], Miss Amy Trebes MSc [98], Mr George MacIntyre-Cockett BSc [98], Alec Birchley MSc [69], Alexander Adams BSc [69], Amy Plimmer [69], Bree Gatica-Wilcox MPhil [69], Dr Caoimhe McKerr PhD [69], Ember Hilvers MA [69], Hannah Jones [69], Dr Hibo Asad PhD [69], Jason Coombes BSc [69], Johnathan M Evans MSc [69], Laia Fina [69], Lauren Gilbert A-Levels [69], Lee Graham BSc [69], Michelle

Cronin [69], Sara Kumziene-SummerhaYes MSc [69], Sarah Taylor [69], Sophie Jones MSc [69], Miss Danielle C Groves BA [93], Mrs Peijun Zhang MSc [93], Miss Marta Gallis MSc [93] and Miss Stavroula F Louka MSc [93].

**Samples and logistics, and Software and analysis tools:**
Dr Igor Starinskij Msc MRCP [48].

**Sequencing and analysis, and Software and analysis tools:**
Dr Chris J Illingworth PhD [47], Dr Chris Jackson PhD [47], Ms Marina Gourtovaia MSc [99], Gerry Tonkin-Hill [99], Kevin Lewis [99], Dr Jaime M Tovar-Corona PhD [99], Dr Keith James PhD [99], Dr Laura Baxter PhD [94], Dr Mohammad T. Alam PhD [94], Dr Richard J Orton PhD [48], Dr Joseph Hughes PhD [48], Dr Sreenu Vattipally PhD [48], Dr Manon Ragonnet-Cronin PhD [39], Dr Fabricia F. Nascimento PhD [39], Mr David Jorgensen MSc [39], Ms Olivia Boyd MSc [39], Ms Lily Geidelberg MSc [39], Dr Alex E Zarebski PhD [23], Dr Jayna Raghwani PhD [23], Dr Moritz UG Kraemer DPhil [23], Joel Southgate MSc [10, 69], Dr Benjamin B Lindsey MRCP [93] and Mr Timothy M Freeman MPhil [93].

**Software and analysis tools, and Visualisation:**
Jon-Paul Keatley [99], Dr Joshua B Singer PhD [48], Leonardo de Oliveira Martins PhD [70], Dr Corin A Yeats PhD [14], Dr Khalil Abudahab PhD [14, 114], Mr Ben EW Taylor MEng [14, 114] and Mirko Menegazzo [14].

**Leadership and supervision:**
Prof John Danesh [99], Wendy Hogsden MSc [46], Dr Sahar Eldirdiri MBBS MSc FRCPath [21], Mrs Anita Kenyon MSc [21], Dr Jenifer Mason MBBS [43], Mr Trevor I Robinson MSc [43], Prof Alison Holmes MD [38, 103], Dr James Price PhD [38, 103], Prof John A Hartley PhD [82], Dr Tanya Curran PhD [3], Dr Alison E Mather PhD [70], Dr Giri Shankar [69], Dr Rachel Jones [69], Dr Robin Howe [69] and Dr Sian Morgan FRCPath [9].

**Metadata curation:**
Dr Elizabeth Wastenge MD [53], Dr Michael R Chapman PhD [34, 88, 99], Mr Siddharth Mookerjee MPH [38, 103], Dr Rachael Stanley PhD [54], Mrs Wendy Smith [15], Prof Timothy Peto PhD [59], Dr David Eyre PhD [59], Dr Derrick Crook [59], Dr Gabrielle Vernet MBBS [33], Dr Christine Kitchen PhD [10], Huw Gulliver [10], Dr Ian Merrick PhD [10], Prof Martyn Guest PhD [10], Robert Munn BSc [10], Dr Declan T Bradley [63, 72] and Dr Tim Wyatt [63].

**Project administration:**
Dr Charlotte Beaver [99], Luke Foulser [99], Sophie Palmer [88], Carol M Churcher [88], Ellena Brooks MA [88], Kim S Smith [88], Dr Katerina Galai PhD [88], Georgina M McManus BSc [88], Dr Frances Bolt PhD [38, 103], Dr Francesc Coll PhD [19], Lizzie Meadows MA [70], Dr Stephen W Attwood PhD [23], Dr Alisha Davies [69], Elen De Lacy MSc [69], Fatima Downing [69], Sue Edwards [69], Dr Garry P Scarlett PhD [76], Mrs Sarah Jeremiah MSc [83] and Dr Nikki Smith PhD [93].

**Samples and logistics:**
Danielle Leek Bsc [88], Sushmita Sridhar BS [88, 99], Sally Forrest BSc [88], Claire Cormie [88], Harmeet K Gill PhD [88], Joana Dias MSc [88], Ellen E Higginson PhD [88], Mailis Maes MPhil [88], Jamie Young BSc [88], Michelle Wantoch PhD [7], Sanger Covid Team (www.sanger.ac.uk/covid-team) [99], Dorota Jamrozy [99], Stephanie Lo [99], Dr Minal Patel PhD [99], Verity Hill [90], Ms Claire M Bewshea MSc [91], Prof Sian Ellard FRCPath [73, 91], Dr Cressida Auckland FRCPath [73], Dr Ian Harrison [66], Dr Chloe Bishop [66], Dr Vicki Chalker [66], Dr Alex Richter PhD [85], Dr Andrew Beggs PhD [85], Dr Angus Best PhD [86], Dr Benita Percival PhD [86], Dr Jeremy Mirza PhD [86], Dr Oliver Megram PhD [86], Dr Megan Mayhew PhD [86], Dr Liam Crawford PhD [86], Dr Fiona Ashcroft PhD [86], Dr Emma Moles-Garcia PhD [86], Dr Nicola Cumley PhD [86], Mr Richard Hopes [64], Dr Patawee Asamaphan PhD [48], Mr Marc O Niebel MSc [48], Prof Rory N Gunson PhD FRCPath [100], Dr Amanda Bradley PhD [52], Dr Alasdair Maclean PhD [52], Dr Guy Mollett MBChB [52], Dr Rachel Blacow MBChB [52], Mr Paul Bird MSc [16], Mr Thomas Helmer [16], Miss Karlie Fallon [16], Dr Julian Tang [16], Dr

Antony D Hale MBBS [49], Dr Louissa R Macfarlane-Smith PhD [49], Katherine L Harper MBiol [49], Miss Holli Carden MSc [49], Dr Nicholas W Machin MSc [45, 64], Ms Kathryn A Jackson MSc [92], Dr Shazaad S Y Ahmad MSc [45, 64], Dr Ryan P George PhD [45], Dr Lance Turtle PhD MRCP [92], Mrs Elaine O'Toole BSc [43], Mrs Joanne Watts BSc [43], Mrs Cassie Breen BSc [43], Mrs Angela Cowell MSc [43], Ms Adela Alcolea-Medina [32, 96], Ms Themoula Charalampous MSc [12, 42], Amita Patel [11], Dr Lisa J Levett PhD [35], Dr Judith Heaney PhD [35], Dr Aileen Rowan PhD [39], Prof Graham P Taylor DSc [39], Dr Divya Shah PhD [30], Miss Laura Atkinson MSc [30], Mr Jack CD Lee MSc [30], Mr Adam P Westhorpe BSc [82], Dr Riaz Jannoo PhD [82], Dr Helen L Lowe PhD [82], Miss Angeliki Karamani MSc [82], Miss Leah Ensell BSc [82], Mrs Wendy Chatterton MSc [35], Miss Monika Pusok MSc [35], Mrs Ashok Dadrah MSc [75], Miss Amanda Symmonds MSc [75], Dr Graciela Sluga MD/MSC [44], Dr Zoltan Molnar PhD [72], Mr Paul Baker MD [79], Prof Stephen Bonner [79], Ms Sarah Essex [79], Dr Edward Barton MD [56], Ms Debra Padgett BSc [56], Ms Garren Scott BSc [56], Ms Jane Greenaway MSc [57], Dr Brendan AI Payne MD [50], Dr Shirelle Burton-Fanning MD [50], Dr Sheila Waugh MD [50], Dr Veena Raviprakash MD [17], Ms Nicola Sheriff BSc [17], Ms Victoria Blakey BSc [17], ms Lesley-Anne Williams BSc [17], Dr Jonathan Moore MD [27], Ms Susanne Stonehouse BSc [27], Dr Louise Smith [55], Dr Rose K Davidson PhD [89], Dr Luke Bedford [26], Dr Lindsay Coupland PhD [54], Ms Victoria Wright BSc [18], Dr Joseph G Chappell PhD [97], Dr Theocharis Tsoleridis PhD [97], Prof Jonathan Ball PhD [97], Mrs Manjinder Khakh [15], Dr Vicki M Fleming PhD [15], Dr Michelle M Lister PhD [15], Dr Hannah C Howson-Wells PhD [15], Dr Louise Berry [15], Dr Tim Boswell [15], Dr Amelia Joseph [15], Dr Iona Willingham [15], Dr Nichola Duckworth [60], Dr Sarah Walsh [60], Dr Emma Wise PhD [2, 111], Dr Nathan Moore PhD [2, 111], Miss Matilde Mori BSc [2, 108, 111], Dr Nick Cortes MRCP FRCPath [2, 111], Dr Stephen Kidd PhD [2, 111], Dr Rebecca Williams BMBS [33], Laura Gifford MSc [69], Miss Kelly Bicknell [61], Dr Sarah Wyllie [61], Miss Allyson Lloyd [61], Mr Robert Impey MSc [61], Ms Cassandra S Malone MSc [6], Mr Benjamin J Cogger BSc [6], Nick Levene MSc [62], Lynn Monaghan [62], Dr Alexander J Keeley MRCP [93], Dr David G Partridge FRCP FRCPath [78, 93], Dr Mohammad Raza [78, 93], Dr Cariad Evans [78, 93] and Dr Kate Johnson [78, 93].

**Sequencing and analysis:**

Emma Betteridge BSc [99], Ben W Farr BSc [99], Scott Goodwin MSc [99], Dr Michael A Quail PhD [99], Carol Scott [99], Lesley Shirley MSc [99], Scott AJ Thurston BSc [99], Diana Rajan MSc [99], Dr Iraad F Bronner PhD [99], Louise Aigrain PhD [99], Dr Nicholas M Redshaw PhD [99], Dr Stefanie V Lensing PhD [99], Shane McCarthy [99], Alex Makunin [99], Dr Carlos E Balcazar PhD [90], Dr Michael D Gallagher PhD [90], Dr Kathleen A Williamson PhD [90], Thomas D Stanton BSc [90], Ms Michelle L Michelsen BSc [91], Ms Joanna Warwick-Dugdale BSc [91], Dr Robin Manley Ph.D. [91], Ms Audrey Farbos MSc [91], Dr James W Harrison Ph.D. [91], Dr Christine M Sambles Ph.D. [91], Dr David J Studholme PhD. [91], Dr Angie Lackenby [66], Dr Tamyo Mbisa [66], Dr Steven Platt [66], Mr Shahjahan Miah [66], Dr David Bibby [66], Dr Carmen Manso [66], Dr Jonathan Hubb [66], Dr Gavin Dabrera [66], Dr Mary Ramsay [66], Dr Daniel Bradshaw [66], Dr Ulf Schaefer [66], Dr Natalie Groves [66], Dr Eileen Gallagher [66], Dr David Lee [66], Dr David Williams [66], Dr Nicholas Ellaby [66], Hassan Hartman [66], Nikos Manesis [66], Vineet Patel [66], Juan Ledesma [67], Ms Katherine A Twohig [67], Dr Elias Allara [64, 88], Ms Clare Pearson [64, 88], Mr Jeffrey K. J. Cheng MSc [94], Dr Hannah E. Bridgewater PhD [94], Ms Lucy R. Frost BSc [94], Ms Grace Taylor-Joyce BSc [94], Dr Paul E Brown PhD [94], Dr Lily Tong PhD [48], Ms Alice Broos BSc [48], Mr Daniel Mair BSc [48], Mrs Jenna Nichols BSc [48], Dr Stephen N Carmichael PhD [48], Dr Katherine L Smollett PhD [40], Dr Kyriaki Nomikou PhD [48], Dr Elihu Aranday-Cortes PhD/DVM [48], Ms Natasha Johnson BSc [48], Dr Seema Nickbakhsh PhD [48, 68], Dr Edith E Vamos PhD [92], Dr Margaret Hughes PhD [92], Dr Lucille Rainbow PhD [92], Mr Richard Eccles MSc [92], Ms Charlotte Nelson MSc [92], Dr Mark Whitehead PhD [92], Dr Richard Gregory PhD [92], Mr Matthew Gemmell MSc [92], Ms Claudia Wierzbicki BSc [92], Ms Hermione J Webster BSc [92], Ms Chloe L Fisher MSc [28], Mr Adrian W Signell BSc [20], Dr Gilberto Betancor PhD [20], Mr Harry D Wilson BSc [20], Dr Gaia Nebbia PhD FRCPath [12], Dr Flavia Flaviani PhD [31], Mr Alberto C Cerda MSc [96], Ms Tammy V Merrill MSc [96], Rebekah E Wilson MSc [96], Mr Marius Cotic MSc [82], Miss Nadua Bayzid BSc [82], Dr Thomas Thompson PhD [72], Dr Erwan Acheson PhD [72], Prof Steven Rushton PhD [51], Prof Sarah O'Brien PhD [51], David J Baker BEng [70], Steven Rudder [70], Alp Aydin MSci [70], Dr Fei Sang PhD [18], Dr Johnny Debebe PhD [18], Dr Sarah Francois PhD [23], Dr Tetyana I Vasylyeva DPhil [23], Dr Marina Escalera Zamudio PhD [23], Mr Bernardo Gutierrez MSc [23], Dr Angela

Marchbank BSc [10], Joshua Maksimovic FD [9], Karla Spellman FD [9], Kathryn McCluggage Msc [9], Dr Mari Morgan PhD [69], Robert Beer BSc [9], Safiah Afifi BSc [9], Trudy Workman HNC [10], William Fuller BSc [10], Catherine Bresner Bsc [10], Dr Adrienn Angyal PhD [93], Dr Luke R Green PhD [93], Dr Paul J Parsons PhD [93], Miss Rachel M Tucker MSc [93], Dr Rebecca Brown PhD [93] and Mr Max Whiteley PhD [93]

**Software and analysis tools:**
James Bonfield BSc [99], Dr Christoph Puethe [99], Mr Andrew Whitwham BSc [99], Jennifier Liddle [99], Dr Will Rowe PhD [41], Dr Igor Siveroni PhD [39], Dr Thanh Le-Viet PhD [70] and Amy Gaskin MSc [69].

**Visualisation:**
Dr Rob Johnson PhD [39].

**1** Barking, Havering and Redbridge University Hospitals NHS Trust, **2** Basingstoke Hospital, **3** Belfast Health & Social Care Trust, **4** Betsi Cadwaladr University Health Board, **5** Big Data Institute, Nuffield Department of Medicine, University of Oxford, **6** Brighton and Sussex University Hospitals NHS Trust, **7** Cambridge Stem Cell Institute, University of Cambridge, **8** Cambridge University Hospitals NHS Foundation Trust, **9** Cardiff and Vale University Health Board, **10** Cardiff University, **11** Centre for Clinical Infection & Diagnostics Research, St. Thomas' Hospital and Kings College London, **12** Centre for Clinical Infection and Diagnostics Research, Department of Infectious Diseases, Guy's and St Thomas' NHS Foundation Trust, **13** Centre for Enzyme Innovation, University of Portsmouth (PORT), **14** Centre for Genomic Pathogen Surveillance, University of Oxford, **15** Clinical Microbiology Department, Queens Medical Centre, **16** Clinical Microbiology, University Hospitals of Leicester NHS Trust, **17** County Durham and Darlington NHS Foundation Trust, **18** Deep Seq, School of Life Sciences, Queens Medical Centre, University of Nottingham, **19** Department of Infection Biology, Faculty of Infectious & Tropical Diseases, London School of Hygiene & Tropical Medicine, **20** Department of Infectious Diseases, King's College London, **21** Department of Microbiology, Kettering General Hospital, **22** Departments of Infectious Diseases and Microbiology, Cambridge University Hospitals NHS Foundation Trust; Cambridge, UK, **23** Department of Zoology, University of Oxford, **24** Division of Virology, Department of Pathology, University of Cambridge, **25** East Kent Hospitals University NHS Foundation Trust, **26** East Suffolk and North Essex NHS Foundation Trust, **27** Gateshead Health NHS Foundation Trust, **28** Genomics Innovation Unit, Guy's and St. Thomas' NHS Foundation Trust, **29** Gloucestershire Hospitals NHS Foundation Trust, **30** Great Ormond Street Hospital for Children NHS Foundation Trust, **31** Guy's and St. Thomas’ BRC, **32** Guy's and St. Thomas’ Hospitals, **33** Hampshire Hospitals NHS Foundation Trust, **34** Health Data Research UK Cambridge, **35** Health Services Laboratories, **36** Heartlands Hospital, Birmingham, **37** Hub for Biotechnology in the Built Environment, Northumbria University, **38** Imperial College Hospitals NHS Trust, **39** Imperial College London, **40** Institute of Biodiversity, Animal Health & Comparative Medicine, **41** Institute of Microbiology and Infection, University of Birmingham, **42** King's College London, **43** Liverpool Clinical Laboratories, **44** Maidstone and Tunbridge Wells NHS Trust, **45** Manchester University NHS Foundation Trust, **46** Microbiology Department, Wye Valley NHS Trust, Hereford, **47** MRC Biostatistics Unit, University of Cambridge, **48** MRC-University of Glasgow Centre for Virus Research, **49** National Infection Service, PHE and Leeds Teaching Hospitals Trust, **50** Newcastle Hospitals NHS Foundation Trust, **51** Newcastle University, **52** NHS Greater Glasgow and Clyde, **53** NHS Lothian, **54** Norfolk and Norwich University Hospital, **55** Norfolk County Council, **56** North Cumbria Integrated Care NHS Foundation Trust, **57** North Tees and Hartlepool NHS Foundation Trust, **58** Northumbria University, **59** Oxford University Hospitals NHS Foundation Trust, **60** PathLinks, Northern Lincolnshire & Goole NHS Foundation Trust, **61** Portsmouth Hospitals University NHS Trust, **62** Princess Alexandra Hospital Microbiology Dept., **63** Public Health Agency, **64** Public Health England, **65** Public Health England, Clinical Microbiology and Public Health Laboratory, Cambridge, UK, **66** Public Health England, Colindale, **67** Public Health England, Colindale, **68** Public Health Scotland, **69** Public Health Wales NHS Trust, **70** Quadram Institute Bioscience, **71** Queen Elizabeth Hospital, **72** Queen's University Belfast, **73** Royal Devon and Exeter NHS Foundation Trust, **74** Royal Free NHS Trust, **75** Sandwell and West Birmingham NHS Trust, **76** School of Biological Sciences, University of Portsmouth (PORT), **77** School of Pharmacy and Biomedical Sciences, University of Portsmouth (PORT), **78** Sheffield Teaching Hospitals, **79** South Tees Hospitals NHS Foundation Trust, **80** Swansea University, **81** University Hospitals Southampton NHS Foundation Trust, **82** University College London, **83** University Hospital Southampton NHS Foundation Trust, **84** University Hospitals Coventry and Warwickshire, **85** University of Birmingham, **86** University of Birmingham Turnkey Laboratory, **87** University of Brighton, **88** University of Cambridge, **89** University of East Anglia, **90** University of Edinburgh, **91**

University of Exeter, **92** University of Liverpool, **93** University of Sheffield, **94** University of Warwick, **95** University of Cambridge, **96** Viapath, Guy's and St Thomas' NHS Foundation Trust, and King's College Hospital NHS Foundation Trust, **97** Virology, School of Life Sciences, Queens Medical Centre, University of Nottingham, **98** Wellcome Centre for Human Genetics, Nuffield Department of Medicine, University of Oxford, **99** Wellcome Sanger Institute, **100** West of Scotland Specialist Virology Centre, NHS Greater Glasgow and Clyde, **101** Department of Medicine, University of Cambridge, **102** Ministry of Health, Sri Lanka, **103** NIHR Health Protection Research Unit in HCAI and AMR, Imperial College London, **104** North West London Pathology, **105** NU-OMICS, Northumbria University, **106** University of Kent, **107** University of Oxford, **108** University of Southampton, **109** University of Southampton School of Health Sciences, **110** University of Southampton School of Medicine, **111** University of Surrey, **112** Warwick Medical School and Institute of Precision Diagnostics, Pathology, UHCW NHS Trust, **113** Wellcome Africa Health Research Institute Durban and **114** Wellcome Genome Campus.
